# *In vivo* demonstration of microvascular thrombosis in severe Covid-19

**DOI:** 10.1101/2020.07.09.20149971

**Authors:** Douglas Alexandre do Espírito Santo, Anna Cristina Bertoldi Lemos, Carlos Henrique Miranda

**Affiliations:** Division of Emergency Medicine, Department of Internal Medicine, Ribeirão Preto School of Medicine, São Paulo University, Ribeirão Preto, SP, Brazil

**Author notes:** **Corresponding author** Carlos Henrique Miranda, MD, PhD, Department of Internal Medicine, Division of Emergency Medicine – São Paulo University, Rua Bernardino de Campos, 1000, Ribeirão Preto – SP 14020-670 – Brazil.

**Keywords:** COVID-19, microvascular thrombosis, sublingual microcirculation, coagulopathy

## Abstract

Several autopsies studies showed the presence of microthrombi in the pulmonary circulation of the severe COVID-19. The major limitation of these investigations is that the autopsy provided static information. Some of these alterations could be secondary to the disseminated intravascular coagulation (DIC) observed as the final common pathway of the multisystem organ failure exhibited in the critical patient. We report the preliminary results of an in *vivo* evaluation of the sublingual microcirculation in thirteen patients with severe COVID-19 requiring mechanical ventilation at the beginning of the hospitalization. They did not have any laboratorial DIC evidence. We observed multiple filling defects moving within the sublingual microvessels indicative of microthrombi in 11 (85%) patients. This is the first imaging documentation of microvascular thrombosis in living patients with severe COVID-19. The clinical relevance of microvascular thrombosis in this disease requires further research.

**Highlights:**

- The presence of microthrombi in the pulmonary circulation is a common finding in autopsies of severe COVID-19.
- In vivo evaluation of the sublingual microcirculation of severe COVID-19 showed multiple filling defects moving within the microvessels indicative of thrombi in most of these patients.
- This is the first imaging documentation of microvascular thrombosis in living patients with severe COVID-19.
- The clinical relevance of microvascular thrombosis in COVID-19 requires further research.

## Introduction

The main clinical presentation of patients with severe coronavirus disease 2019 (COVID-19) is an acute respiratory failure with significant hypoxemia, which requires mechanical ventilation [1]. In addition to the gas exchange involved, recently published investigations described a high incidence of thrombotic events in these patients [2].

Several autopsies studies demonstrated the presence of microthrombi in the pulmonary circulation [3-6]. The major limitation of these investigations is that the autopsy provided static information. Some of these alterations could be secondary to the disseminated intravascular coagulation (DIC) observed as the final common pathway of the multisystem organ failure exhibited in the critical patient.

To support the microvascular thrombosis occurrence in severe COVID-19 pathophysiology, we report the preliminary results of an *in vivo* evaluation of the sublingual microcirculation allowing the dynamic visualization of these microvessels.

## Methods

We included adult patients with SARS-Cov-19 virus infection confirmed through reverse transcriptase-polymerase chain reaction (RT-PCR) of nasopharyngeal swabs and severe clinical presentation with respiratory failure requiring mechanical ventilation. The sublingual microcirculation visualization was performed with the patient in dorsal decubitus with the headboard raised to 45 degrees. We used the Capiscope HVCS handheld video capillaroscopy system (KK Technology, Honiton, England, UK), and imaging was analyzed using the Glycocheck system (Microvascular health solution, American Fork, UT, USA). The camera, protected with disposable tips, was placed under the tongue without pressing this instrument in at least five different regions inside the mouth after the patient’s admission into the intensive care unit.

The ethical board of our institution approved this investigation. The relatives of the patients gave their written informed consent to participate in this research.

## Results

We analyzed the imaging of the sublingual microcirculation of thirteen patients on the first day after hospital admission. The demographics, clinical, and laboratory characteristics of these patients are presented in Table 1. All the patients were on mechanical ventilation, and they did not have any evidence of circulatory shock. Six patients (46%) were receiving a low norepinephrine dose to support sedation and neuromuscular blocking agents, both of them with normal lactate. None of these patients met the criteria for DIC. Despite elevated D-dimer levels, prothrombin time, and activated partial thromboplastin time (aPTT) were normal, fibrinogen levels were increased in all patients, and mild thrombocytopenia was observed in only two patients.

**Table 1.** Characteristics of the COVID-19 patients in whom sublingual microcirculation was assessed.

| Parameters | n=13 |
| --- | --- |
| Demographic |  |
| Age (years), mean±sd | 58±11 |
| Male, n.(%) | 10(77) |
| Clinical presentation |  |
| Fever, n.(%) | 11(85) |
| Cough, n.(%) | 12(92) |
| Dyspnea, n.(%) | 13(100) |
| Myalgia, n.(%) | 7(54) |
| Duration of symptoms (days), median (IQR) | 6(4-9) |
| Risk factors |  |
| Diabetes mellitus, n.(%) | 4(31) |
| Hypertension, n.(%) | 4(31) |
| Cardiovascular disease, n.(%) | 1(08) |
| Immunocompromise, n.(%) | 1(08) |
| BMI (Kg/m<sup>2</sup>), mean±sd | 34±9 |
| Physical Examination |  |
| Systolic blood pressure (mmHg), mean±sd | 120±23 |
| Diastolic blood pressure (mmHg), mean±sd | 74±11 |
| Heart rate (beats per minute), mean±sd | 79±19 |
| Mechanical ventilation, n.(%) | 13(100) |
| Tidal volume (ml), mean±sd | 374±66 |
| PEEP (cm of water), mean±sd | 12±3 |
| Plateau pressure (cm of water), mean±sd | 24±4 |
| Static compliance (ml/cm of water), mean±sd | 33±8 |
| Respiratory rate (cycles/min), mean±sd | 24±4 |
| Fio<sub>2</sub> (%), range (min-max) | 0.55-1.00 |
| Drugs |  |
| Norepinephrine, n.(%) | 6(46) |
| Midazolam, n.(%) | 13(100) |
| Fentanyl, n.(%) | 13(100) |
| Neuromuscular blocking agent, n.(%) | 13(100) |
| Laboratory test |  |
| Hemoglobin (g/dl), mean±sd | 13±2 |
| White-cell count (per microliter), mean±sd | 8232±4149 |
| Platelets count (per microliter), mean±sd | 234231±55853 |
| Creatinine clearance (ml/min), mean±sd | 69±18 |
| Creatinine (mg/dl), mean±sd | 1.2±0.3 |
| D-dimer (µg/L), median (IQR) | 1830(1120-2320) |
| Fibrinogen (mg/dl), median (IQR) | 767(672-843) |
| aPTT (Ratio), median (IQR) | 1.16(1.06-1.20) |
| Prothrombin Time (INR), median (IQR) | 1.06(1.03-1.18) |
| C-reactive protein (mg/L), mean±sd | 18±8 |
| Lactate (mg/dl), mean±sd | 1.7±0.4 |
| Pao<sub>2</sub>/Fio<sub>2</sub> relation, mean±sd | 128±32 |
| Chest radiography with bilateral opacities, n.(%) | 13(100) |
| Scores, median (IQR) |  |
| SOFA | 10(8-11) |
| SAPS 3 | 56(49-68) |
| DIC score (ISTH) | 2(2-3) |
| SIC score | 2(2-2) |
| ARDS classification |  |
| Severe, n.(%) | 2(15) |
| Moderate, n.(%) | 11(85) |

sd: standard deviation; IQR: interquartile range; BMI: body mass index; PEEP: positive end-expiratory pressure; Fio<sub>2</sub>: fraction of inspired oxygen; aPTT: activated partial thromboplastin time; INR: international normalized ratio; SOFA: sequential organ failure assessment score; SAPS 3: simplified acute physiology score 3; DIC: International Society of Thrombosis and Haemostasis criteria for disseminated intravascular coagulation; SIC: sepsis-induced coagulopathy score; ARDS: acute respiratory distress syndrome

We observed some evidence of microvascular thrombosis in 11 (85%) patients. For comparison, an example of the normal sublingual microcirculation in a healthy individual is presented in Figures 1A and 1B. The arrows in Figures 1C, 1D, and 1E illustrate multiple filling defects moving within the microvessels indicative of microthrombi, which were found in 11 (85%) patients. Four (31%) patients exhibited complete stagnated capillaries. Microvessels with the interrupted flow and semi-oval imaging in its distal extremity suggest acute thromboembolic occlusion (asterisk in Figure 1F) in 5 (38%) patients. An abruptly thromboembolic obstruction of a microvessel was registered during the evaluation in 3 patients (videos in supplementary material).

**Figure 1.**
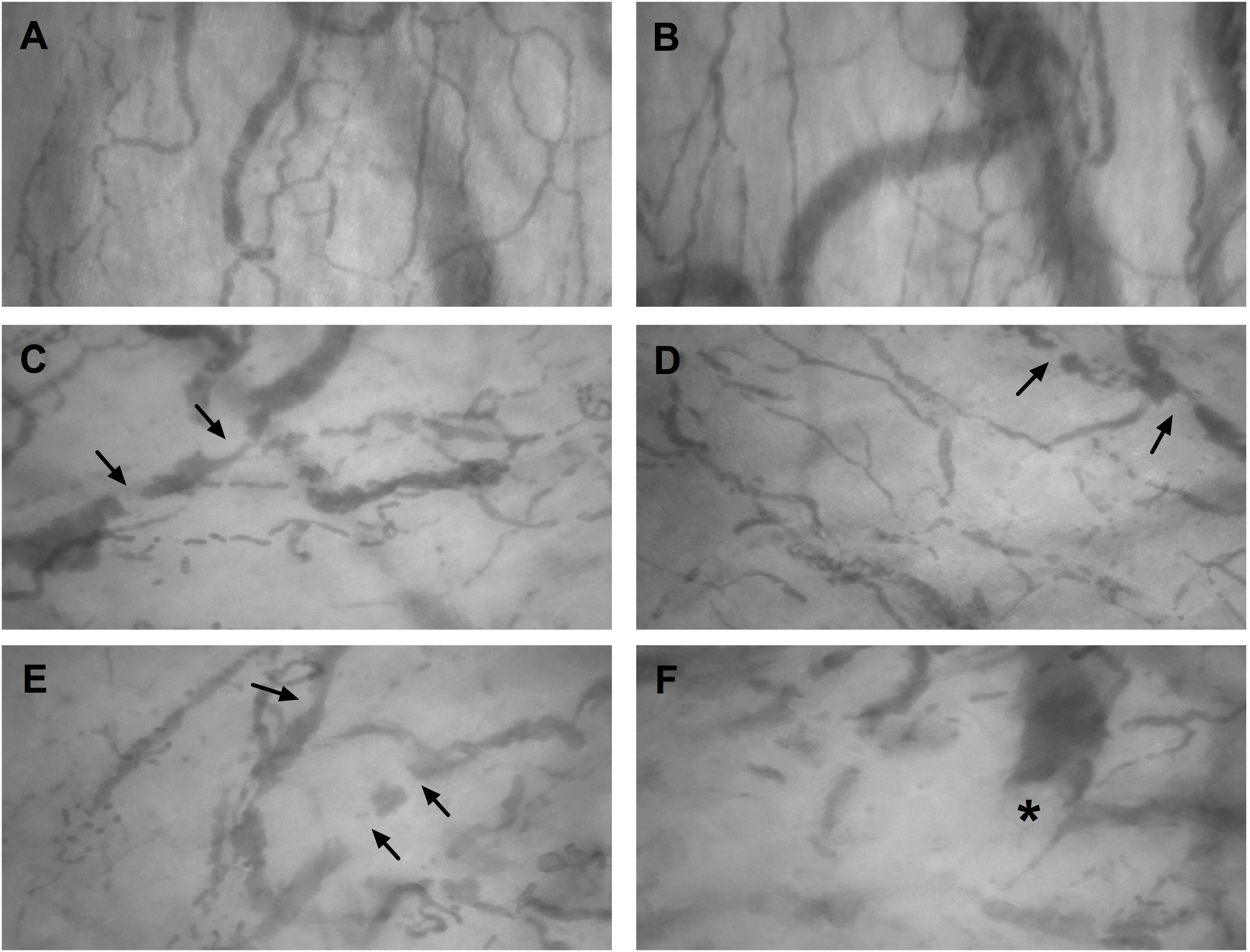
Normal sublingual microcirculation in a healthy individual (A-B). Presence of several filling defects (arrows) moving inside the sublingual microvessels indicative of microthrombi (C-E). Microvessel with interrupted flow showing semi-oval imaging (asterisk) in its distal extremity compatible with acute thromboembolic occlusion (F).

## Discussion

Thrombus in small vessels of the pulmonary circulation is a common finding in the autopsies of COVID-19 patients. Dolhnikoff et al. [3] observed a variable number of small fibrinous thrombi in small pulmonary arterioles in areas of both damaged and more preserved lung parenchyma in 8 out of 10 cases in the minimally invasive autopsy. Small fibrinous thrombi were rarely found in the glomeruli and superficial dermal vessels. Ackermann et al. [4] examined seven lungs obtained during the autopsy and found widespread pulmonary vessels’ thrombosis. Fox et al. [5] observed the presence of thrombosis and microangiopathy in the lungs’ small vessels in autopsies of ten African American decedents. Carsana et al. [6] analyzed lung tissue samples of 38 patients, besides the diffuse alveolar damage, they observed fibrin thrombi in the small arterial vessels in 87% of cases, around half of which had more involvement than 25% of the lung tissue.

Our investigation demonstrated thrombi in microcirculation since the beginning of the hospitalization, and none of these patients had criteria for DIC. This finding reinforces that microvascular thrombosis is a hallmark of the COVID-19.

Another conclusion emphasized by our investigation is that microvascular thrombosis occurs systemically, and it could affect different organs. Probably those organs with a high capillary density, such as the lungs, are the most compromised. In the lung, pulmonary microvascular thrombosis could lead to a dead space effect (ventilated areas, but not perfused), contributing to the critical hypoxemia observed in these cases. However, microvascular thrombosis could compromise other areas such as the kidney, liver, and brain, contributing to multiple organ dysfunction, such as recent descriptions [7]. This mechanism could explain the findings of Tang et al. [8], who in a retrospective study achieved a low 28-day mortality in severe Covid-19 patients with the use of anticoagulant therapy (heparin) for seven days or more, especially the ones with high sepsis-induced coagulopathy score (≥ 4) or high D-dimer (≥3.0 mg/l).

This virus causes an intense inflammatory response with massive pro-inflammatory cytokines release, such as interleukin (IL)-1, IL-6, IL-8, and interferon-γ, associated with endothelial vascular damage causing a hypercoagulability status, which leads to microvascular thrombosis [9,10]. Some authors suggest using MicroCLOTS (microvascular COVID-19 lung vessels obstructive thromboinflammatory syndrome) as an adequate name for this severe pulmonary presentation [11]. As all these alterations are diffuse, we expected to find systemic microvascular thrombosis, as we described in the sublingual territory. Since the pulmonary capillaries assessment is challenging in living patients, we generalize that the same should happen in the pulmonary microcirculation.

This is a descriptive study of a small series of COVID-19 patients. There was no control group in this investigation; however, normally these findings are not observed in the sublingual microcirculation of healthy individuals. In summary, this is the first imaging documentation of microvascular thrombosis in living patients with severe COVID-19 since the beginning of the hospitalization. The clinical relevance of microvascular thrombosis in COVID-19 requires further research.

## Data Availability

The data used and/or analyzed during the current study are available from the corresponding author on reasonable request.

## Declarations

### Ethical approval

The study was approved by the Research Ethics Committee of our institution and followed the Declaration of Helsinki.

### Consent to participate

The subjects or their relatives gave their written informed consent to participate in this research.

### Consent for publication

Not applicable.

### Author’s contributions

DAES-acquisition of data, analysis and interpretation of data, draft the article, final approval of the last version. ACBL-acquisition of data, analysis and interpretation of data, draft the article, final approval of the last version. CHM-conception and design of the study, acquisition of data, analysis and interpretation of data, draft the article, final approval of the last version.

